# Characteristics of Cardiac Memory in Patients with Implanted Cardioverter Defibrillator: the CAMI study

**DOI:** 10.1101/19005181

**Authors:** Kazi T. Haq, Jian Cao, Larisa G. Tereshchenko

## Abstract

**Objective:** The goal of this study was to determine factors associated with cardiac memory (CM) in patients with implantable cardioverter-defibrillators (ICD).

**Methods:** Patients with structural heart disease (n=20; mean age 72.6±11.6 y; 80% male; mean left ventricular ejection fraction (LVEF) 31.7±7.6%; history of myocardial infarction (MI) in 75%, ventricular tachycardia (VT) in 85%) and preserved atrioventricular (AV) conduction received primary (80%) or secondary (20%) prevention dual-chamber ICD. Standard 12-lead ECG was recorded in AAI and DDD mode, before and after 7 days of right ventricular (RV) pacing in DDD mode with short AV delay. Direction (azimuth and elevation) and magnitude of spatial QRS, T, and ventricular gradient (SVG) vectors were measured before and after 7 days of RV pacing. CM was quantified as the degree of alignment between QRS_DDD-7_ and T_AAI-7_ vectors (QRS_DDD-7_-T_AAI-7_ angle). Circular statistics and mixed models with a random slope and intercept were adjusted for days 1-7 change in cardiac activation, LVEF, known risk factors, and use of medications known to affect CM.

**Results:** QRS_DDD-7_-T_AAI-7_ angle strongly correlated (circular r = - 0.972; P<0.0001) with T_AAI-7_-T_DDD-7_ angle. In the mixed models, history of MI (−180°(95%CI −320° to −40°); P=0.011) and female sex (−162°(95%CI −268° to −55°); P=0.003) counteracted CM-T azimuth changes (+132°(95%CI 80°-184°); P<0.0001). History of VT (+27(95%CI 4-46) mV*ms; P=0.007) amplified CM-T area increase (+15(95%CI 6–24) mV*ms; P<0.0001).

**Conclusions:** Existing cardiac remodeling affects CM in response to RV pacing. Women develop less CM than men. Activation memory is another manifestation of CM.

---

Right ventricular (RV) apical pacing can cause pacing-induced cardiomyopathy^1, 2^: nearly every fifth patient develops pacing-induced cardiomyopathy if RV pacing burden ≥20%^3, 4^. Biventricular pacing and His bundle pacing^5^ are more physiological, but also more technically challenging pacing approaches. An informed clinical decision regarding the choice of pacing approach should be based on the evaluation of risks and benefits for each patient. Unfortunately, it remains largely unknown which factors (besides pacing burden) are associated with pacing-induced cardiomyopathy.

RV pacing changes activation pathway and induces complex repolarization phenomenon of cardiac memory (CM).^6^ CM is a form of cardiac remodeling caused by altered myocardial stretch.^7^ Altered ventricular stretch and subsequent local cardiac angiotensin II release but not altered ventricular activation initiates CM.^8^ The T wave changes of CM result from underlying changes in ion channels (*I*_to_, *I*_kr_, *I*_Ca,L_) and connexin43 remodeling.^7^ CM can be fully reversible after return to normal activation pathway. All these facts suggest that the amount of CM developed in response to RV pacing is likely associated with clinical outcomes. However, while molecular mechanisms of CM have been extensively studied,^6^ its translation into clinical practice has been limited.^9^ Current clinical perception of CM is restricted by recognition of T wave inversion that develops after a period of altered ventricular activation once normal ventricular activation is restored, differentiating CM from ischemic T wave inversion.^10^

Previously, CM was studied in patients undergoing pacemaker implantation.^11-14^ An interaction of RV pacing-induced CM with preexisting cardiac remodeling [e.g. after myocardial infarction (MI) and ventricular tachycardia (VT)] remains incompletely understood. The goal of this study was to determine factors associated with CM in patients receiving implantable cardioverter defibrillators (ICD). We hypothesized that preexisting cardiac remodeling is associated with CM.

## Methods

The MATLAB (MathWorks, Inc, Natick, MA) software code for ECG data analysis is provided at https://physionet.org/physiotools/globalelectricalheterogeneity/, https://github.com/Tereshchenkolab/Origin, and https://github.com/Tereshchenkolab/cardiacmemory. In order to minimize the possibility of unintentionally sharing information that can be used to re-identify private information, a subset of the data generated for this study are available at the GitHub and can be accessed at https://github.com/Tereshchenkolab/cardiacmemory/QRS_T_DDD2_AAI2.

### Study population

The Cardiac Memory with ICD (CAMI) prospective study was conducted by Medtronic. The study participants were enrolled at the Beth Israel Deaconess Medical Center (BIDMC), and the data analysis was performed at the Oregon Health & Science University (OHSU). The study was approved by the Institutional Review Boards at the BIDMC and the OHSU. All study participants signed an informed consent before entering the study.

The CAMI study included adults above 18 years of age who have received a Medtronic market released dual-chamber ICD with chronically implanted (for at least 3 months, ≥ 90 days) Medtronic RV leads with SVC coil and RV ring electrode (Sprint Fidelis, Sprint Quattro Secure 6947, Sprint Quattro 6944, etc.) for approved indications. The RV tip location must be in the RV apex. Patients must have sinus rhythm with 1:1 AV conduction at physiological heart rates at baseline.

Exclusion criteria were: (1) a history of unstable angina pectoris within the last 3 months unless treated by coronary intervention; (2) inability to tolerate DDD pacing or AAI pacing due to subjective discomfort, heart failure, or other reasons; (3) anti-tachycardia pacing or shock therapy from ICD for spontaneous tachyarrhythmia episodes for the last 3 months; (4) more than 1% of RV pacing for the last 3 months (confirmed by the ICD device interrogation); (5) NYHA class III-IV congestive heart failure; (6) left ventricular ejection fraction (LVEF) < 20%; (7) baseline ECG abnormalities (complete left bundle branch block, T wave inversions secondary to left ventricular hypertrophy) precluding expression of CM; (8) inaccessible for follow-up at the study center.

### ECG recording and pacing protocol: induction of cardiac memory

At the baseline study visit, resting supine 12 lead electrocardiogram (ECG) was recorded using a MAC 5000 electrocardiograph (GE Marquette, Milwaukee, WI), in AAI and DDD mode with a short AV delay at a rate 10% faster than presented sinus rhythm. Then, ICD devices were programmed in the DDD mode with a short (100-120 ms) paced and sensed AV delay and lower rate as clinically indicated.

The second study visit was conducted after seven days of ventricular pacing when ICD devices were interrogated, and the percentage of ventricular pacing data was collected. During the follow-up visit, resting 12-lead ECG was recorded first in DDD mode, and then in AAI mode. The pacing rate during ECG recording on a follow-up visit maintained the same as at the baseline ECG recording.

### Measurement of cardiac memory on the body surface vectorcardiogram

The raw digital 12-lead ECG signal (sampling rate 500Hz, amplitude resolution 1µV) was analyzed at OHSU. Kors matrix^15^ was used to transform the 12-lead ECG into orthogonal XYZ ECG. All 10-second ECG recordings were reviewed, and all beats were manually labeled (LGT). Ectopic beats, fusion beats, and artifact-distorted beats were excluded from the analysis. Four types of median beats were constructed: AAI mode atrial-paced ventricular-sensed (APVS) beat recorded on the study day one (AAI-1) and day seven (AAI-7), and DDD mode atrial-paced ventricular-paced (APVP) beat recorded on the study day one (DDD-1) and day seven (DDD-7). The heart vector origin point was identified as previously described.^16^ Fiducial points (QRS onset and offset, and T offset) were automatically detected on a vector magnitude. Accuracy of the origin point and fiducial point detection was verified using a visual aid (KTH, LGT). Spatial peak and area QRS, T, and spatial ventricular gradient (SVG) vectors were defined as previously described,^17^ and their direction (azimuth and elevation) and magnitude were measured. The scalar value of SVG was measured by the sum absolute QRST integral (SAI QRST)^18, 19^ and QT integral on vector magnitude signal (iVMQT).^17^

CM was quantified after seven days of ventricular pacing, as the degree of alignment between VP QRS vector (QRS_DDD-7_) and VS T vector (T_AAI-7_), measured as QRS_DDD-7_-T_AAI-7_ angle. Changes in ventricular repolarization were assessed as T_AAI-1_-T_AAI-7_ and T_DDD-1_-T_DDD-7_, as well as T_AAI-1_-T_DDD-1_ and T_AAI-7_-T_DDD-7_ angles.

The difference in ventricular activation between VS and VP QRS vectors was measured by QRS_AAI-7_-QRS_DDD-7_ angle. To eliminate an error due to possible differences in ECG leads placement between days one and seven, CM angles were measured on the same day recordings. To assess a possible error due to ECG leads placement on two different days, we measured spatial angles QRS_DDD-1_-QRS_DDD-7_ and QRS_AAI-1_-QRS_AAI-7_, and an agreement between angles QRS_AAI-7_-QRS_DDD-7_ and QRS_AAI-1_-QRS_DDD-1_.

### Ventricular pacing vector

On VCG, VP vector was defined as a median VP spike calculated on the 10-sec recording.

### Statistical analyses

The distribution of all variables was evaluated. Normally distributed continuous variables were presented as mean ± standard deviation (SD). A paired *t*-test was used to compare normally distributed VCG parameters at different pacing modes, at baseline and after seven days of ventricular pacing.

Circular statistics were used to analyze circular variables (spatial angles, azimuth, and elevation). To describe circular variable, mean circular direction and 95% confidence interval (CI) were reported. Non-uniformity of the circular variable distribution was confirmed by the Rayleigh test and the Kuiper test for all studied circular variables. A paired comparison of circular variables was performed using the Hotelling’s paired test. The circular-circular correlation coefficients between two circular variables were calculated by the Fisher & Lee method. The circular-linear correlation coefficients were calculated by the Fisher, Mardia & Jupp method. The Watson U-square statistic and the Kuiper statistics were used for two-sample tests for circular variables. To account for multiple testing in correlation analyses, the correlation was considered statistically significant if the P-value was less than 0.001.

To determine associations of demographic and clinical characteristics with changes in T and QRS vectors over the course of 7 days, we conducted longitudinal analysis and constructed two sets of mixed models. One set of models was built to predict changes in direction and magnitude of T area vector, separately in the AAI (VS) mode and DDD (VP) mode. Another set of models was built to predict changes in the direction and magnitude of the QRS area vector in DDD (VP) mode. As there was prominent person-to-person variability in QRS and T vector changes, we constructed mixed models with a random slope and intercept. The Hausman specification test confirmed the consistency of random effect estimates for all models. We used an unstructured covariance structure. A likelihood ratio test confirmed a better model fit for a random slope, for all models. To test our hypothesis that preexisted remodeling can affect the development of CM, we constructed two models. Model 1 was adjusted for age, sex, and two major causes of preexisted CM: a history of MI and VT. Model 2 was in addition adjusted for other known factors affecting cardiac remodeling: LVEF, history of diabetes, hypertension, use of angiotensin-converting enzyme inhibitors (ACEi) or angiotensin receptor blockers (ARBs), and class III antiarrhythmic (AA) drugs. To adjust for possible unmeasured confounders (e.g. due to differences in ECG leads location in days 1 and 7, or unmeasured disease-related factors), both models 1 and 2 were adjusted for longitudinal change in corresponding QRS variable. Model of T (and SVG) azimuth change was adjusted for QRS azimuth change. Model of T (and SVG) elevation change was adjusted for QRS elevation change. The model of T area change was adjusted for the QRS area change. In addition, we tested the hypothesis that T azimuth (and T area) change during abnormal activation (in DDD mode) is associated with QRS azimuth (and QRS area) change, adjusting for the same confounders in models 1 and 2. In mixed models, a P value less than 0.05 was considered significant.

STATA MP 16 (StataCorp LP, College Station, TX) and Oriana-Circular Statistics 4 (Kovach Computing Services, Pentraeth, Wales, UK) were used for statistical analyses.

## Results

### Study population

The clinical characteristics of the study participants are shown in Table 1. Most of the study participants were men with ischemic cardiomyopathy and ICD implanted for primary prevention of sudden cardiac death (SCD). Of note, 85% had a history of VT, and nearly half had a sustained VT. The vast majority of participants were on beta-blockers and ACEi/ARBs, and one-third of participants received class III antiarrhythmic medications (sotalol or amiodarone). During the seven study days, all study participants experienced constant RV pacing: the average percentage of RV pacing was 99.73±0.23% (range 99.3-100%).

**Table 1.** Clinical characteristics of study participants

| Characteristic | All participants (n=20) |
| --- | --- |
| Age(SD), y | 72.6(11.1) |
| Female, n(%) | 4 (20) |
| Percentage of ventricular pacing during 7 days in the study(SD) | 99.7(0.2) |
| LVEF(SD), % | 31.7(7.6) |
| Primary prevention ICD, n(%) | 16(80) |
| NYHA class II-III, n(%) | 10(50) |
| History of coronary heart disease, n(%) | 17(85) |
| History of myocardial infarction, n(%) | 15(75) |
| Location of infarction: anterior, n(% of patients with MI Hx) | 4(27) |
| Infero-posterior, n(% of patients with MI Hx) | 9(60) |
| Multiple (inferior and anterior, anterior and posterolateral) | 2(13) |
| History of revascularization, n(%) | 13(65) |
| Hypertension, n(%) | 10(50) |
| Diabetes, n(%) | 7(35) |
| Atrial fibrillation, n(%) | 9(45) |
| History of ventricular tachycardia, n(%) | 17(85) |
| History of sustained ventricular tachycardia, n(%) | 8(40) |
| Class III antiarrhythmic medications, n(%) | 6(30) |
| Beta-blockers, n(%) | 19(95) |
| Digoxin, n(%) | 4(20) |
| Calcium channel blocker, n(%) | 2(10) |
| Diuretics, n(%) | 9(45%) |
| ACE inhibitors / angiotensin receptor blockers, n(%) | 16(80) |

ACE=angiotensin converting enzyme

### Development of cardiac memory

Mean CM angle (*µ*) was 67.5° (95%CI 47.6° – 87.4°); median 58.2°; length of mean vector *r* was 0.727; concentration (κ) was 2.2; circular variance 0.27; circular SD 45.7°.

In paired comparison, there were no differences in RR’ interval across all four recordings (Table 2). At baseline, in APVS AAI-1 beat, the QRS vector pointed to the left and slightly backward, whereas the T vector pointed straight forward (indicating preceding remodeling), resulting in a wide baseline QRS-T angle.

**Table 2.**
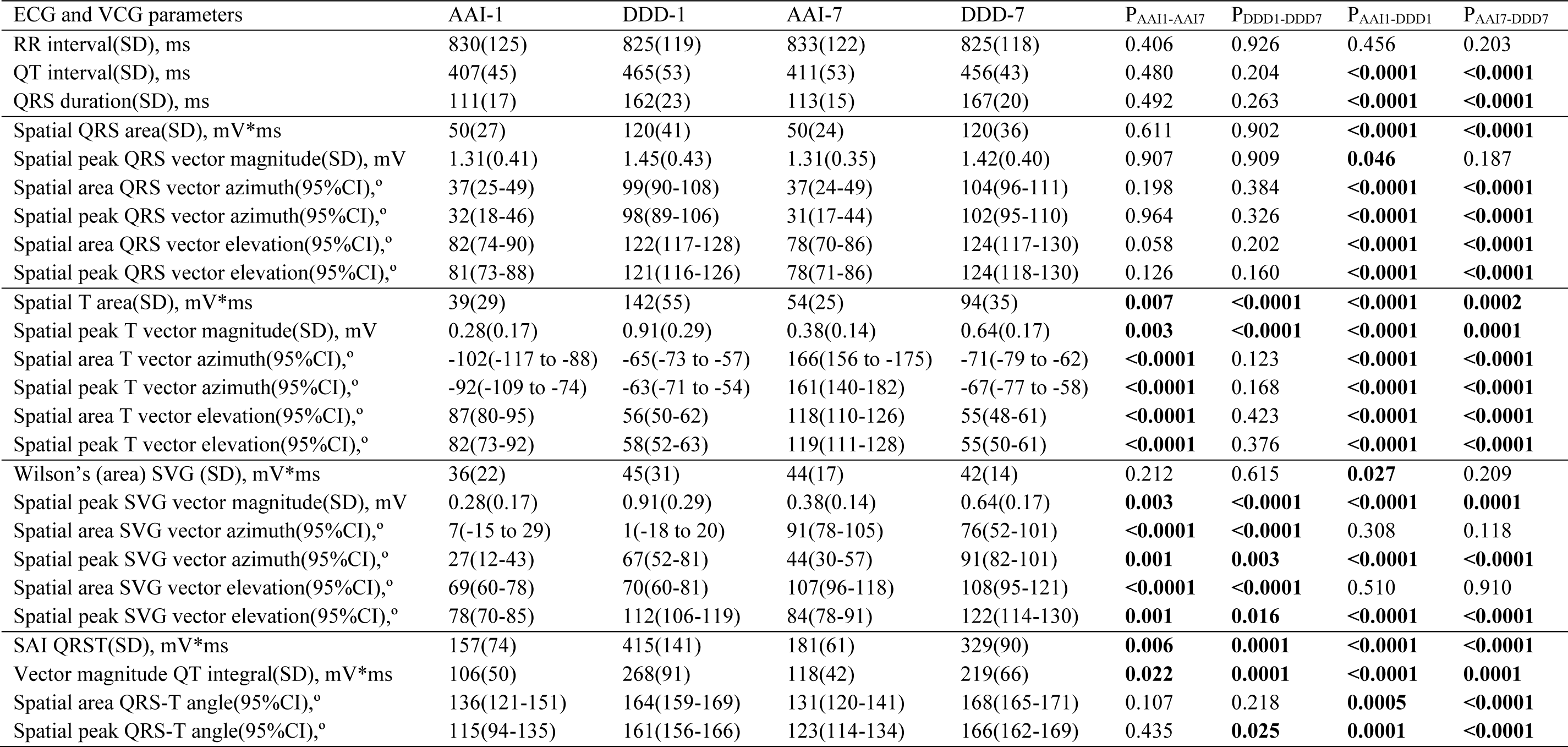
Cardiac memory development

Ventricular pacing in day one (APVP DDD-1 beat) caused QT prolongation, QRS widening, rotation of QRS vector further backward and up, rotation of T vector down and leftward, and further widening of QRS-T angle. Notably, the direction of Wilson’s SVG vector did not change, whereas its magnitude slightly increased. Magnitudes of QRS and T vectors and areas also increased (Table 2).

After seven days of VP and return to normal activation (APVS AAI-7 beat), the QRS vector returned to the same direction as in APVS AAI-1 beat. QT and QRS intervals, QRS-T angle, and SVG magnitude in AAI-7 did not differ from AAI-1. As expected, CM on APVS beat manifested by prominent T vector magnitude enlargement, increased SVG and SAI QRST, and dramatic changes in T vector direction (turned sharply to the right and upward), and SVG vector direction (turned upward and backward).

On the 7^th^ day, we observed very similar differences between APVS and APVP beats (AAI-7 vs. DDD-7), as on the first study day. Neither magnitude nor direction of Wilson’s SVG differed between APVS and APVP beats.

Comparison of APVP DDD-1 and DDD-7 beats revealed no differences in direction and magnitude of the QRS vector, the direction of the T vector, and the QRS-T angle. However, the direction of the SVG vector changed dramatically (turned upward and backward), and magnitudes of T, SVG, and SAI QRST significantly decreased. The development of CM is shown in Figure 1.

**Figure 1.**
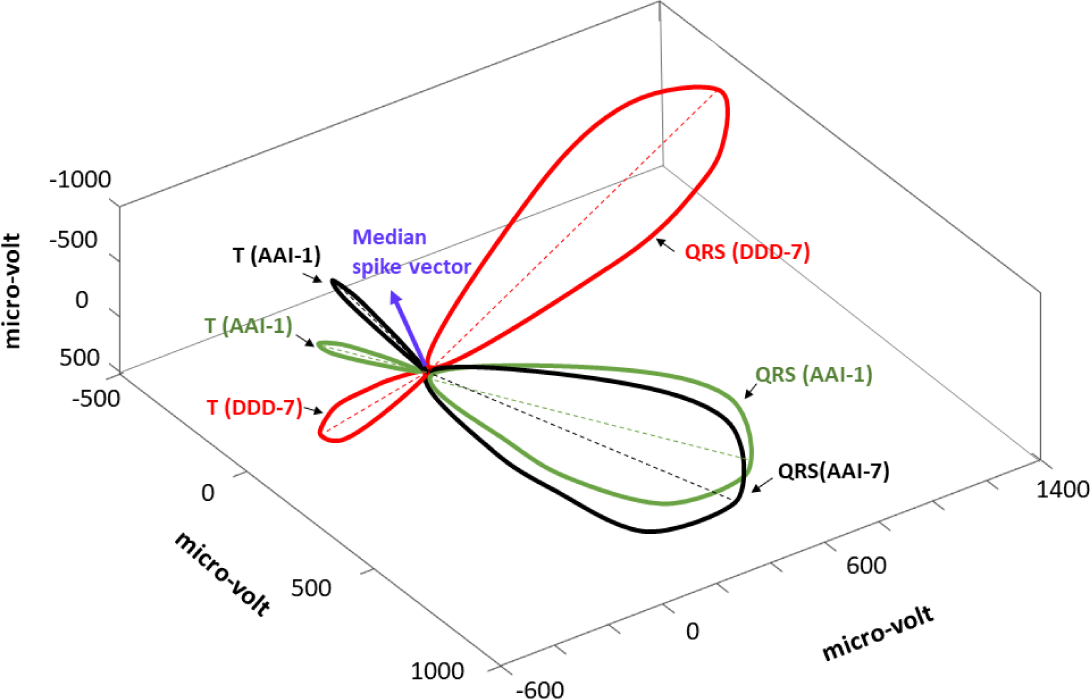
A representative example of QRS and T loops with corresponding peak vectors in median beats recorded at baseline (AAI-1; green) and after the development of CM (AAI-7; black, and DDD-7; red). Median pacing spike vector is shown as a purple arrow.

### Baseline repolarization characteristics associated with cardiac memory

CM angle perfectly (r = - 0.972) correlated with T_AAI-7_-T_DDD-7_ angle that reflects difference in repolarization in two different activation patterns after development of CM (Figure 2). Correlations between CM (QRS_DDD-7_-T_AAI-7_) angle and T_AAI-7_-T_AAI-7_ (r = - 0.198), T_DDD-1_-T_DDD-7_ (r = - 0.081), and T_AAI-1_-T_DDD-1_ (r = - 0.373) angles was weak and non-significant.

**Figure 2.**
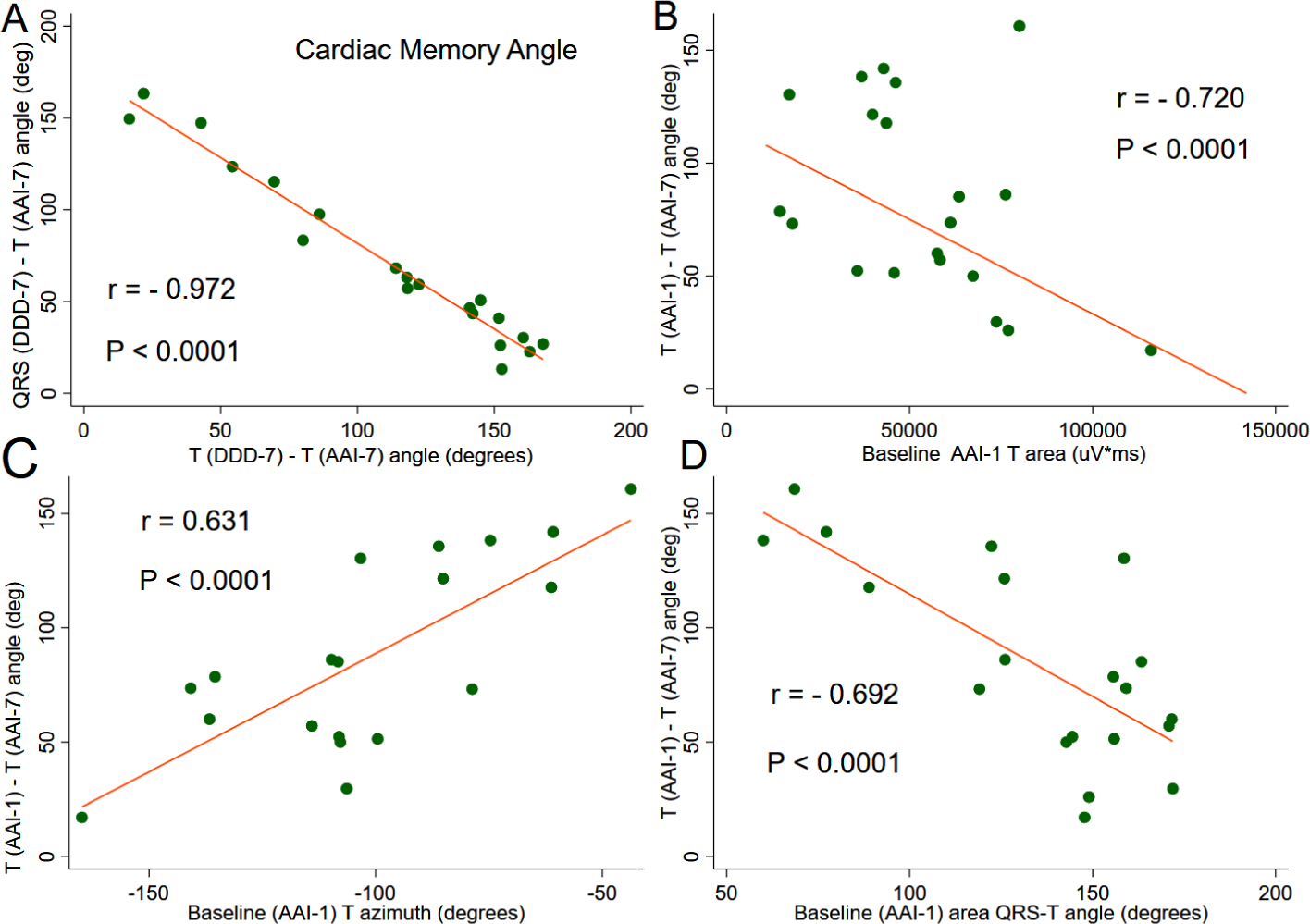
Repolarization memory. (**A**) Scatterplot of QRS_DDD-7_ – T_AAI-7_ angle (y-axis) against T_DDD-7_ – T_AAI-7_ angle (x-axis). Scatterplot of T_DDD-7_ – T_AAI-7_ angle (y-axis) against (**B**) T_AAI-1_ area (x-axis), (**C**) T_AAI-1_ azimuth (x-axis), and (**D**) QRS-T_AAI-1_ angle. A linear best fit line is shown.

T_AAI-1_-T_AAI-7_ angle negatively correlated with baseline T_AAI-1_ peak magnitude (r = - 0.738; P<0.0001) and T _AAI-1_ area (r = - 0.720; P<0.0001), and positively correlated with T _AAI-1_ azimuth (Figure 2), both T _AAI-1_ peak azimuth (r = 0.623; P<0.0001) and T _AAI-1_ area azimuth (r = 0.631; P<0.0001). Baseline QRS-T _AAI-1_ angle negatively correlated with T_AAI-1_-T_AAI-7_ angle (r = - 0.692; P<0.0001; Figure 2).

### Baseline characteristics associated with ventricular activation pattern during RV pacing

The difference in ventricular activation between VS and VP QRS vectors as measured by QRS_AAI-7_-QRS_DDD-7_ angle was on average 77.4°; length of mean vector 0.927; median 82.6° (95%CI 67.7° – 87.2°). In the paired analysis, there was no difference between QRS_AAI-7_-QRS_DDD-7_ and QRS_AAI-1_-QRS_DDD-1_ angles. A possible error due to variations in ECG electrodes placement was below 10 degrees. Mean QRS_AAI-1_-QRS_AAI-7_ angle was 7.5°; length of mean vector 0.997; median 6.5° (95%CI 5.5°-9.5°). Mean QRS_DDD-1_-QRS_DDD-7_ angle was 9.6°; length of mean vector 0.976; median 5.0 (95%CI 4.1°-15.1°).

We observed a significant correlation between baseline APVP (DDD-1) repolarization characteristics and differences in ventricular activation in AAI and DDD mode after CM has been developed, as measured by QRS_AAI-7_-QRS_DDD-7_ angle. Baseline APVP T_DDD-1_ azimuth correlated with QRS_AAI-7_-QRS_DDD-7_ angle (r = 0.604; P<0.001; Figure 3), but not QRS_AAI-1_-QRS_DDD-1_ angle (r = 0.355; NS). Baseline APVP QT_DDD-1_ interval was moderately strongly correlated with APVP QRS_DDD-7_ azimuth (r = 0.623; P<0.001; Figure 3), whereas the correlation of APVS QT_AAI-1_ interval with APVP QRS_DDD-7_ azimuth was weak (r = 0.436; P<0.05). Baseline APVS T_AAI-1_ vector magnitude (r = 0.653) and T_AAI-1_ area (r = 0.601) positively correlated with QRS_DDD-7_ vector elevation on the 7^th^ day during ventricular pacing (Figure 3), but not in a normal ventricular conduction (QRS_AAI-7_ vector elevation: r = 0.261 for T_AAI-1_ vector magnitude and r = 0.333 for T_AAI-1_ area).

**Figure 3.**
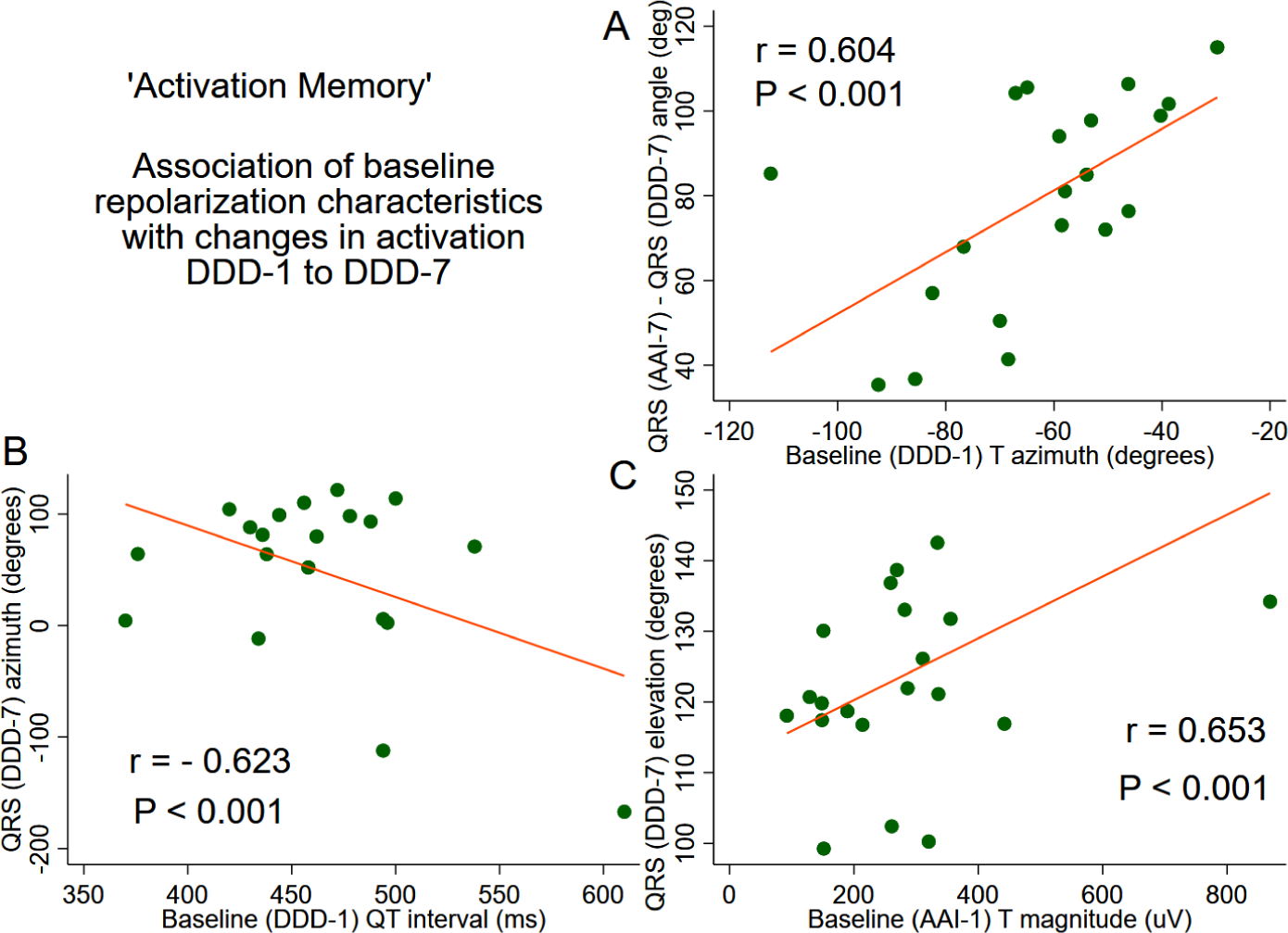
Activation memory. (A) Scatterplot of QRS_AAI-7_ – QRS_DDD-7_ angle (y-axis) against T_DDD-1_ azimuth. (B) Scatterplot of QRS_DDD-7_ azimuth (y-axis) against QT_DDD-1_ interval. (C) Scatterplot of QRS_DDD-7_ elevation against T_AAI-1_ magnitude. A linear best fit is shown.

### The direction of Ventricular Pacing vector

VP vector was directed upward and rightward, consistently with implanted RV lead. In the paired analysis, there was no difference in VP vector azimuth recorded on day one versus day 7 (165°; (95%CI 157°-173°) vs. 173; (95%CI 157°-189°); P=0.099). VP vector elevation slightly decreased by 10° one week after RV lead implantation: V _DDD-1_ elevation 110°; (95%CI 102°-117°) vs. V _DDD-7_ elevation 100° (95%CI 91°-110°); P=0.006. There were no meaningful correlations of VP vector direction with cardiac activation or repolarization metrics.

### Clinical characteristics associated with cardiac memory

We observed prominent person-to-person variability in changes of T and SVG vectors that manifest CM (Figure 4). Mixed models analyses results showed that in AAI mode, T azimuth displayed the most dramatic changes from day 1 to day 7 (Table 3). History of MI and female sex was associated with the significant opposite effect on T azimuth, counteracting the development of CM. Change in T elevation was associated only with a change in QRS elevation, but not any clinical or demographic characteristics. History of MI, history of VT, and history of class III antiarrhythmics use was associated with T magnitude changes (Table 3). In DDD mode, the history of VT was associated with changes in SVG azimuth during seven days of follow-up. History of MI, VT, and female sex were associated with changes in SVG elevation, reducing the degree of CM manifestation (Table 3). Similarly, female sex and VT history were associated with T magnitude changes in DDD mode, offsetting the development of CM.

**Table 3.**
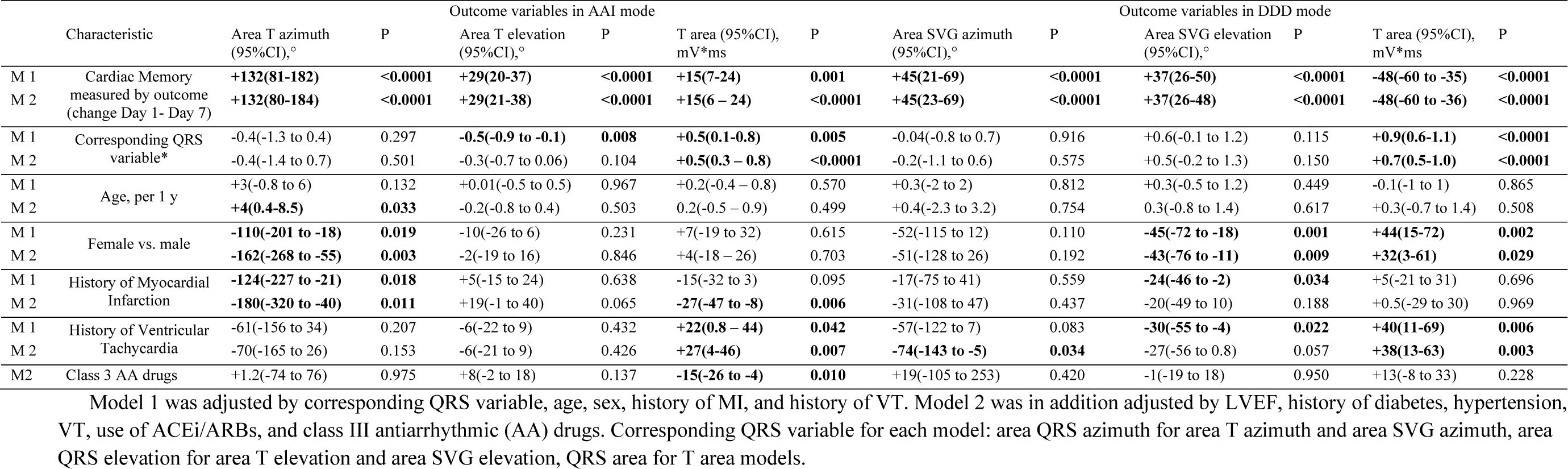
Demographic and clinical characteristics associated with cardiac memory

**Figure 4.**
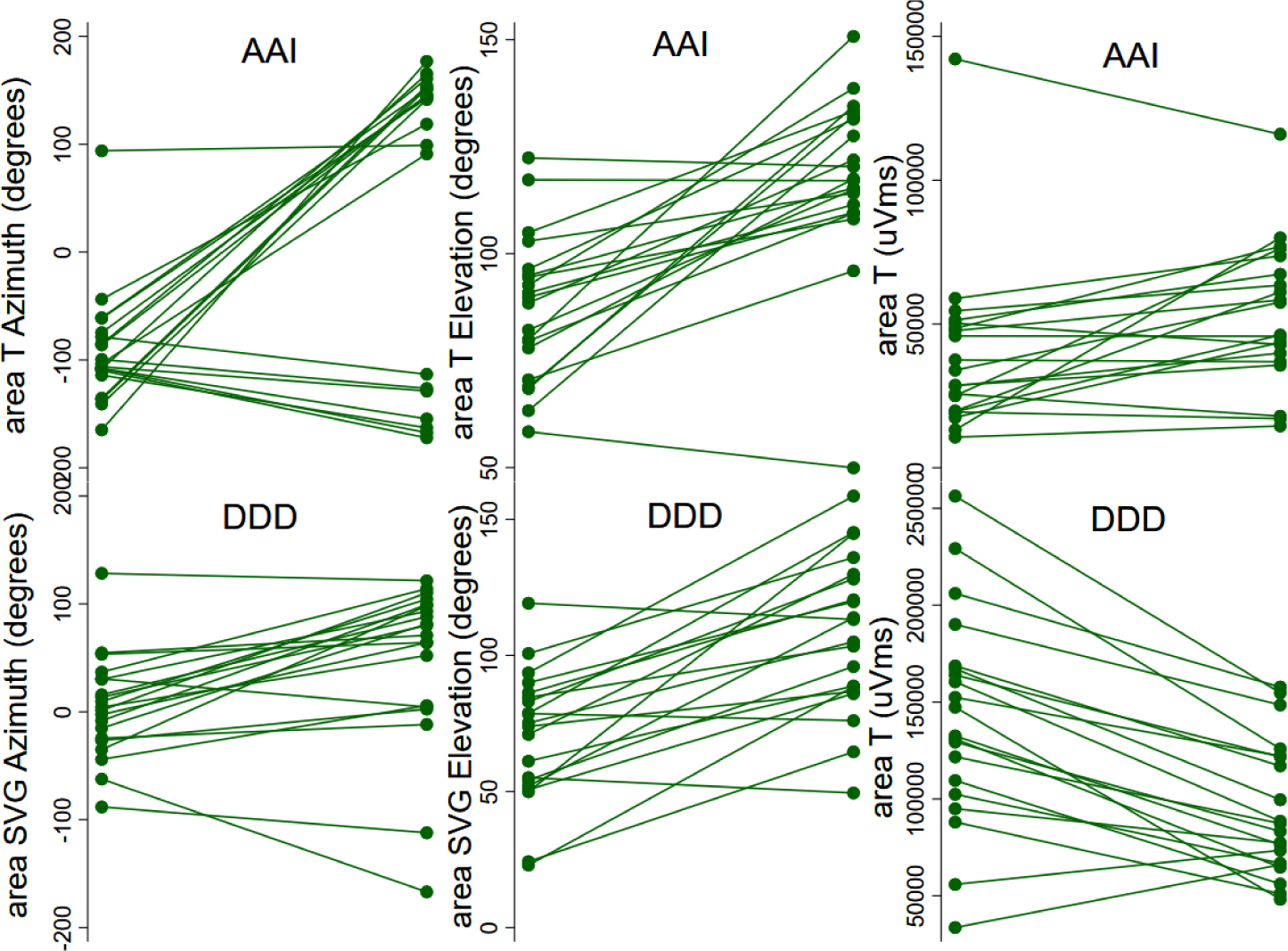
A manifestation of cardiac memory in AAI and DDD mode. Time-series line plots show the change in repolarization from day 1 to day 7, for every study participant. Top panels show the change in T azimuth, T elevation, and area T in AAI mode. The bottom panel shows the change in SVG azimuth, SVG elevation, and area T in DDD mode.

In an adjusted mixed model analysis, we detected changes in ventricular activation as a manifestation of CM. In DDD mode, we observed significant changes in QRS azimuth and area, from day 1 to day 7 (Table 4). Female sex and history of MI were associated with a reduction of QRS azimuth changes, whereas diabetes and LVEF worsening were associated with greater QRS azimuth changes. Similar associations were observed for the QRS area. In AAI mode, there were no statistically significant changes in QRS vector direction and magnitude from day 1 to day 7.

**Table 4.**
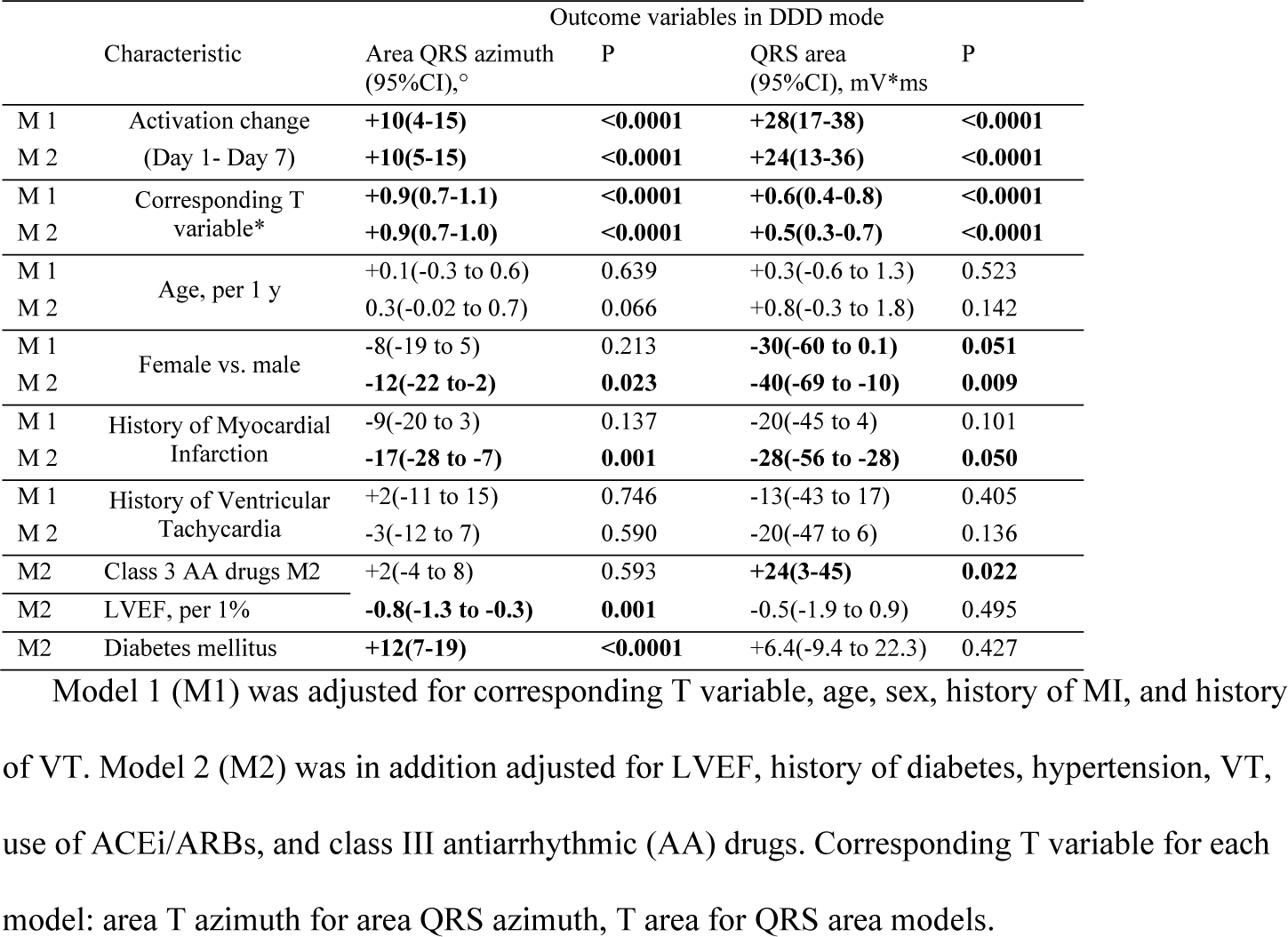
Demographic and clinical characteristics associated with change in activation (QRS vector) in DDD mode

## Discussion

In this prospective study of CM in ICD patients, we observed several novel findings. First, we showed that existing cardiac remodeling due to MI and VT significantly affects the degree of CM in response to RV pacing. Secondly, we, for the first time, demonstrated sex differences in CM development. After adjustment for the type of cardiomyopathy, degree of LV dysfunction, use of medications, and major cardiovascular risk factors (hypertension and diabetes), women developed less CM as compared to men. Thirdly, we noticed that in participants with preceded cardiac remodeling, the CM is associated with significant changes in cardiac activation. While repolarization memory “remembers” abnormal activation after baseline activation has been restored, activation memory “remembers” normal baseline activation during abnormal (VP) activation, resulting in a smaller angle between QRS vectors of normal (baseline) and abnormal (VP) activation. Further studies of repolarization and activation memory are needed to understand the mechanisms and clinical significance of CM.

### Cardiac remodeling is associated with the degree of repolarization memory

We observed that the more abnormal baseline repolarization was, the less repolarization memory had been developed in response to ventricular pacing. The wider the baseline spatial QRS-T angle was, the less repolarization memory was developed. The larger baseline T area was, the less repolarization memory was developed. Similarly, the more abnormal was the direction of the baseline T vector, the less repolarization memory was developed. The abnormal (rightward-forward) direction of T vector, large magnitude of T, and wide QRS-T angle are well-known signs of earlier cardiac memory or cardiac remodeling. A more negative T vector azimuth in CM patients with DDD RV pacing^20^ is consistent with our finding of less repolarization memory with more negative T vector azimuth (Figure 2). Accordingly, in the mixed model analyses adjusted for a change in cardiac activation between days 1 and 7, LV systolic function, known risk factors, and use of medications known to affect the development of cardiac memory, history of MI and VT strongly counteracted repolarization changes. History of MI and VT nearly completely canceled manifestation of CM, both in AAI and DDD modes. Thus, the robust development of typical CM in response to RV pacing suggests an absence or a minimal degree of preexisted cardiac remodeling. In contrast, weak repolarization response to RV pacing implies that repolarization ion channels in ventricular cardiomyocytes had been already remodeled, as shown in many *in vitro* studies,^7, 21, 22^ and display a saturation of response. Our finding is consistent with CM studies in HF patients undergoing bi-ventricular pacing.^12, 13^

### Women develop less cardiac memory than men

Previous studies using animal models^22-24^ or human subjects,^6, 9, 11^ did not investigate sex differences in CM. Interestingly, in our study, sex was strongly associated with the degree of CM manifestation. In fully adjusted mixed models, female sex was strongly and independently associated with a significantly smaller amount of CM, suggesting that women developed less dyssynchrony in response to RV pacing, as compared to men. Female sex nearly completely canceled manifestation of CM, both in AAI and DDD modes (Table 3). The finding of sex differences in CM is novel and has an important clinical significance. It is well known that women benefit from cardiac resynchronization therapy (CRT) more than men.^25^ We speculate that sex differences in CM contribute to sex differences in CRT response.

### “Activation memory” is another manifestation of cardiac memory

A large number of previous studies described CM as a repolarization phenotype.^6, 9, 11, 21, 24, 26^ In this study, we, for the first time described the manifestation of activation memory. In addition to repolarization memory, which manifests by the wide angle between T vectors in AAI and DDD modes on the 7^th^ day, activation memory is revealed by the narrow angle between QRS vectors in AAI and DDD modes on the 7^th^ day. Repolarization memory manifests by T_AAI-7_ vector aligning with abnormal activation vector; repolarization “remembers” abnormal activation. In turn, activation memory manifests during continued abnormal activation, by abnormal activation vector aligning with normal activation vector. Activation memory is likely a compensatory mechanism attempting to minimize dyssynchrony developing in response to sustained abnormal activation. Activation and repolarization are two phases of the repeated cycle, following one after another. Development of repolarization memory affects refractoriness in ventricles, which, in turn, affects the way how ventricles activate. Further studies of activation memory mechanisms and its clinical significance are needed.

### Spatial ventricular gradient reflects cardiac memory

Our results one more time^27^ confirmed Wilson’s ventricular gradient^28^ concept, suggesting that the SVG is largely independent of the ventricular activation sequence (no difference between SVG_AAI-7_ and SVG_DDD-7_).^29, 30^ SVG is determined by the heterogeneity in the whole area under the action potential across the heart, rather than by heterogeneity in the action potential duration alone.^29-31^ SVG vector tracked the development of CM better than the T vector (Table 2), especially in DDD mode. Thus, change in SVG direction can be used to assess CM in persistently abnormal activation (during continuous ventricular pacing, bundle branch block).

### Limitations

The study size was small. Validation of the study findings in a larger study is required.

## Data Availability

The software code for the electrocardiogram and vectorcardiogram data analysis is provided as open-source.

https://github.com/Tereshchenkolab/cardiacmemory

https://github.com/Tereshchenkolab/Origin

https://physionet.org/physiotools/geh/

---

**Supplemental Table 1.** Peak vectors measurements of cardiac memory development

| ECG and VCG parameters | AAI-1 | DDD-1 | AAI-7 | DDD-7 | P<sub>AAI1-AAI7</sub> | P<sub>DDD1-DDD7</sub> | P<sub>AAI1-DDD1</sub> | P<sub>AAI7-DDD7</sub> |
| --- | --- | --- | --- | --- | --- | --- | --- | --- |
| Spatial peak QRS vector magnitude(SD), mV | 1.31(0.41) | 1.45(0.43) | 1.31(0.35) | 1.42(0.40) | 0.907 | 0.909 | **0.046** | 0.187 |
| Spatial peak QRS vector azimuth(95%CI),° | 32(18-46) | 98(89-106) | 31(17-44) | 102(95-110) | 0.964 | 0.326 | **<0.0001** | **<0.0001** |
| Spatial peak QRS vector elevation(95%CI),° | 81(73-88) | 121(116-126) | 78(71-86) | 124(118-130) | 0.126 | 0.160 | **<0.0001** | **<0.0001** |
| Spatial peak T vector magnitude(SD), mV | 0.28(0.17) | 0.91(0.29) | 0.38(0.14) | 0.64(0.17) | **0.003** | **<0.0001** | **<0.0001** | **0.0001** |
| Spatial peak T vector azimuth(95%CI),° | -92(-109 to -74) | -63(-71 to -54) | 161(140-182) | -67(-77 to -58) | **<0.0001** | 0.168 | **<0.0001** | **<0.0001** |
| Spatial peak T vector elevation(95%CI),° | 82(73-92) | 58(52-63) | 119(111-128) | 55(50-61) | **<0.0001** | 0.376 | **<0.0001** | **<0.0001** |
| Spatial peak SVG vector magnitude(SD), mV | 0.28(0.17) | 0.91(0.29) | 0.38(0.14) | 0.64(0.17) | **0.003** | **<0.0001** | **<0.0001** | **0.0001** |
| Spatial peak SVG vector azimuth(95%CI),° | 27(12-43) | 67(52-81) | 44(30-57) | 91(82-101) | **0.001** | **0.003** | **<0.0001** | **<0.0001** |
| Spatial peak SVG vector elevation(95%CI),° | 78(70-85) | 112(106-119) | 84(78-91) | 122(114-130) | **0.001** | **0.016** | **<0.0001** | **<0.0001** |
| Spatial peak QRS-T angle(95%CI),° | 115(94-135) | 161(156-166) | 123(114-134) | 166(162-169) | 0.435 | **0.025** | **0.0001** | **<0.0001** |

